# Computed Tomography 3D Segmentation Based on Gray-Level Histograms

**DOI:** 10.1101/2022.08.26.22279285

**Authors:** Carlos T. Guimeráns-Otero, Fernando Martín-Rodríguez, Fernando Isasi-de-Vicente, Mónica Fernández-Barciela

## Abstract

3D imaging technologies like CT (Computed Tomography) and MRI (Magnetic Resonance Imaging) have represented a great advance for diagnosis. These studies are composed of many 2D grayscale images that represent slices of patient’s body, normally orthogonal to body main axis (from head to feet, normally axis Z in Cartesian representation). These slices (normally called axial slices) are good for many diagnosis issues, but sometimes it is interesting to consider the whole study as a 3D volume. This paper is about segmenting 3D volumes obtained from medical studies (mainly CT). We base ourselves on studying the statistical distribution of gray levels so that we can segment different tissues and treating them as separate 3D objects. Note than in a CT image, gray level is basically proportional to tissue density and this technique should be good to distinguish hard tissues like bones or teeth from soft ones like muscles or skin.

## 1. Introduction

Nowadays, medical imaging technologies like CT (Computed Tomography) and MRI (Magnetic Resonance Imaging) [1], provide 3D information of the inner structures of human body. Primarily, this information is obtained in the form of “axial” (XY plane) slices that are 2D gray level images where gray level has some “tissue related” meaning:

- In CT, a big value in gray level is related with a big attenuation for X-Ray signals, which normally means a bigger density.
- In MRI, images are obtained from the response of odd spin atoms (mainly hydrogen) to magnetic field variations. A MRI image can be tuned in different manners to reveal different tissue characteristics (MRI can be weighted to ρ, T_1_ and T_2_ parameters).

Although information comes in the form of slices, together all slices convey 3D information. It is common to stack all slices to get a 3D volume where pixels have evolved into voxels that are averages of some physical characteristic over small cubes (or sometimes rectangular cuboids). From this mathematical structure is easy to extract slices parallel to other planes. Most used are “sagittal” (XZ plane) and “coronal” (YZ plane).

One step further is getting in some manner to classify voxels as pertaining to the same tissue, this would allow to extract 3D objects from the whole volume. These objects can be represented in 3D rendering systems or even 3D/VR displays. Objects information can be exported to 3D CAD file formats to allow other applications, for example: creating 3D models with 3D printing.

This paper is about tissue segmentation using gray level values. Generation of 3D representation using user provided thresholds is a common practice using software like 3D Slicer [2] or InVesalius [3].

Of course, there exist other approaches to tissue segmentation. For example, in summary [4], authors describe segmentations based on other algorithms: “region growing”, “watershed” and “level set”. These methods will be briefly described and compared to ours in the discussion section. Advancing a glimpse, our method is good for CT images and not so useful on MRI (we have not tested it on other image technologies like PET, although it would be possible). These other methods are better on MRI images but are very dependent on proper initialization. In this case, our method could be used to automate this initialization.

All our development was implemented using the MATLAB environment [5] that has a lot of tools to ease image applications.

## 2. Materials and Methods

### 2.1 Histogram Analysis

Histogram is simply the count of the number of pixels for each possible Gray-Level value. Sometimes the horizontal (Gray-Level) axis is divided into a finite number of intervals or bins (256 is a typical value). Sometimes histograms are divided by its own summation (number of pixels) to get an approximation of the probability density function for Gray-Level (understood as a random variable). The histogram of an image gives us information about different significant regions (objects) in that image. The analysis of a histogram can lead to the computation of appropriate thresholds that divide the image into different regions of interest. A classic example is the computation of a threshold to divide between characters and background in a Gray-Level image coming from a document scanner (typical in OCR applications).

Another example, more related with this research, can be given explaining figure 2. In this case, we have a CT study of the jaws/teeth zone. From a single slice image (top left), we compute the histogram (top right). See that intuitively, we have three regions: between 0.0 and 0.1 (Gray Level is normalized between 0.0 and 1.0), we have basically empty regions; between 0.1 and 0.4, we have soft tissues; above 0.4, we have the hard tissues (bone and teeth). The images below are renderings of 3D reconstructions for points between 0.1 and 0.4 (down left, that shows patient’s skin and even some hair) and above 0.4 (down right, that shows skull and teeth).

**Fig. 1.**
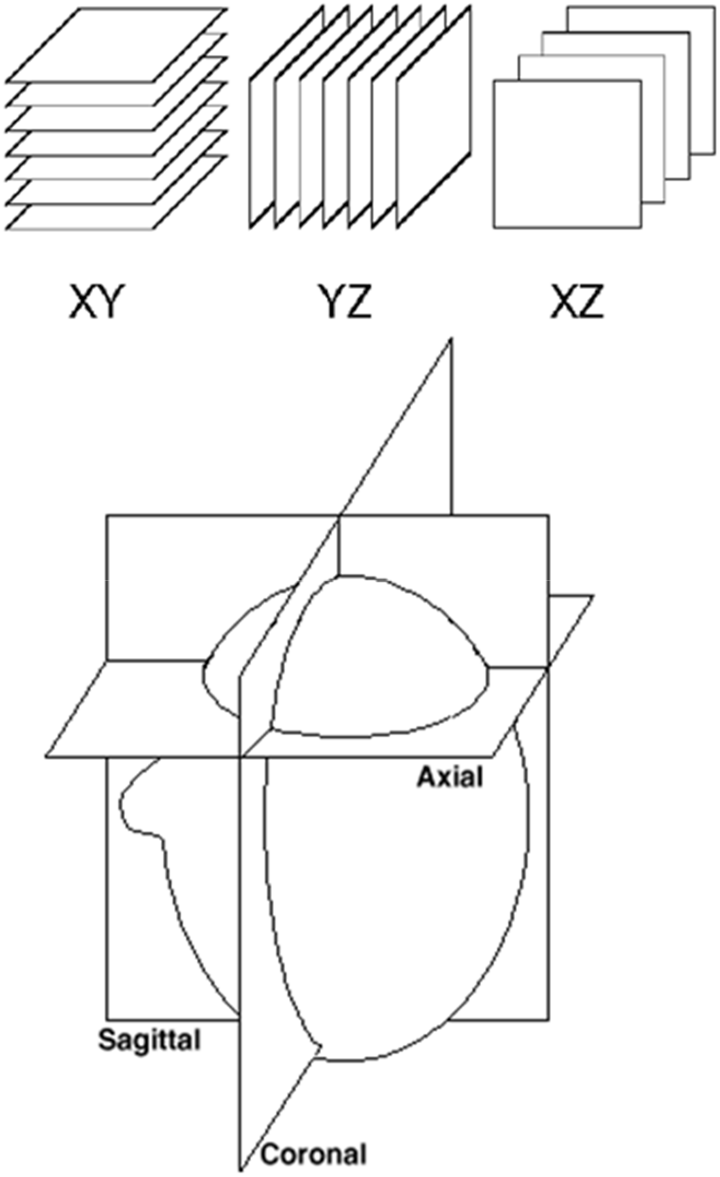
Axial, sagittal and coronal slices.

**Fig. 2.**
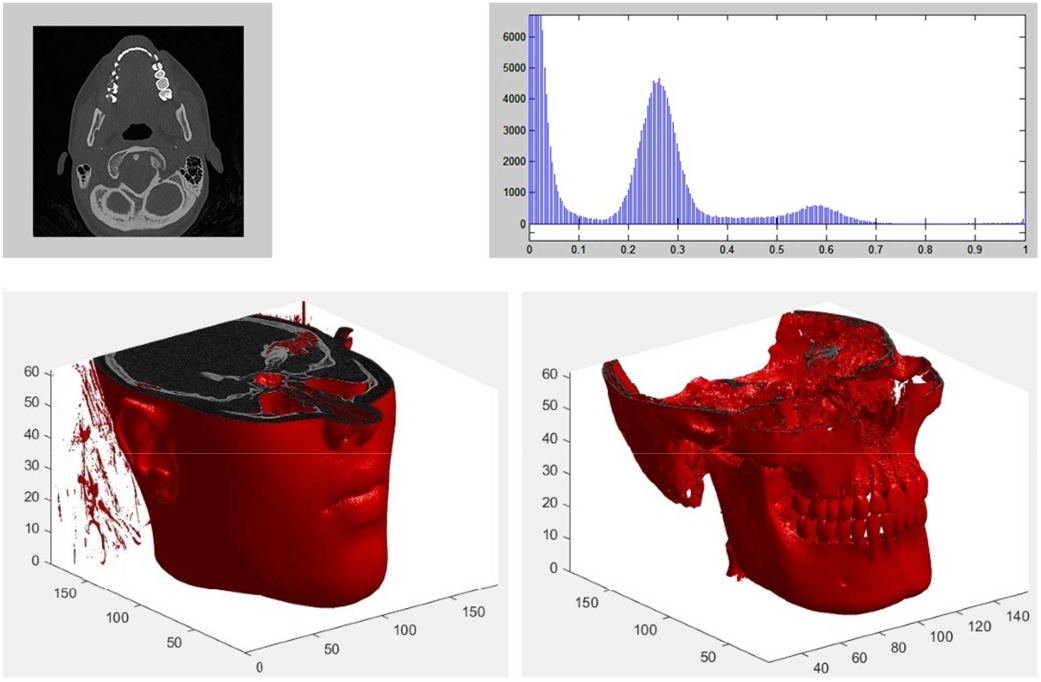
Segmentation example (visual threshold computation).

Note that this research is about the automatic extraction of these threshold values (0.1 and 0.4 in the example).

The problem of finding thresholds for the case of two interest regions (bimodal histograms) has been solved time ago by N. Otsu [6]. According to Otsu, we must maximize a quality function. For each possible threshold T_h_, this number is derived from considering the histogram divided into two marginal probability functions (T_h_ is the frontier between them). The function to maximize is: D = σ^2^ − (w_1_σ^2^_1_ + w _2_ σ^2^_2_), called “between-class variance”; where σ is the global deviation, σ_i_ are partial deviations and w_i_ are the total probabilities of each partial distribution. Equations for D can be rewritten in this manner:

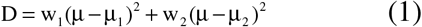

Where μ and μ_i_ are the global and partial means. Equation for D can also be seen as the numerator of the F quotient in an ANOVA study. To compute N thresholds, we will use the methods published in [7]. In this paper, histogram is divided into N+1 marginal distributions. The expression for D in this case would be:

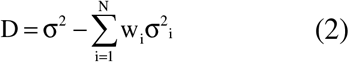

Again this can be rewritten:

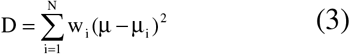

To optimize (3), it would be necessary to test all possible N-tuples of thresholds and we will have a very large number of possibilities. For histograms with 256 bins we would have:

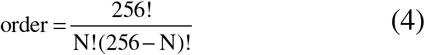

For 3 thresholds (N =3), result is 2.7 million of 3-tuples. This method is really precise but ot has been found to be very slow.

Another method defined in [7] is “peak detection”. This is a method based on heuristically studying the histogram curve. See figure 2, and note that regions can be revealed from their most outstanding points (local maxima in histogram). Thresholds or “frontier points” between two regions will be the local minima between each two local maxima.

Peak detection is based on non-linear histogram filtering. We define a sliding window; a point will be a local maximum when it is the absolute maximum of its window (window centered in the interest point).

### 2.2 Global Histogram

If we have a volume constructed from planar slices, we can speak about a global histogram simply changing the concept of pixel for voxel. This global volume histogram can be computed from the whole volume or can be obtained adding the individual slice histograms (not normalized).

We applied the analysis methods described in the previous section to these histograms getting correct segmentations in simple CT images like that in figure 2. We also noted that for studies of “bigger extension” like a whole thorax CT, global histogram is not appropriate. Best results are got with the “generalized” (multi-threshold) Otsu, if speed of study is required, “peak detection” is preferred.

### 2.3 Localized Histogram

The problems when processing large body parts were due to the fact that in these cases, a set of unique thresholds for the whole study does not exist. For this reason, we developed a more general method that consists of dividing the whole study into small sets of slices. A “local” histogram is computed for each subset and a local set of thresholds is computed using “peak detection”.

### 2.4 Segmenting Objects

After computing histograms, we can get 3D representations of different tissues. A step further would be to extract separate 3d structures corresponding to anatomic objects: bones, arteries…

To get this complex issue, we use the fact that 3D representation in MATLAB is based in the function **isosurface** [8] that converts the point cloud into a mesh of faces (triangles) with known vertices. Applying an algorithm called splitFV [9], we can group together the contiguous faces obtaining separate 3D objects.

### 2.5 Exporting Volumes

Last step is exporting the segmented objects into a 3D file format that can be read by CAD (or even 3D printing/prototyping applications). Using free tools [10], we managed to export isosurface objects into STL file format [11] that is accepted for all interesting 3D CAD applications.

## 3. Results

We have applied the described method (local histogram + analysis by peak detection), we have obtained good results for CT images. Test images have been downloaded from public sites as: www.aycan.de/lp/sample-dicom-images.html, https://www.dicomlibrary.com/, https://www.kaggle.com/search?q=dicom+in%3Adatasets.

Some examples of software execution are summarized in the following images:

As it was explained in the previous sections, objects can be separately exported into STl files to be loaded and managed by CAD applications. As an example, we exported a model from the aorta in figure 3 and we opened the resulting file with a common 3D application: Blender.

**Fig. 3.**
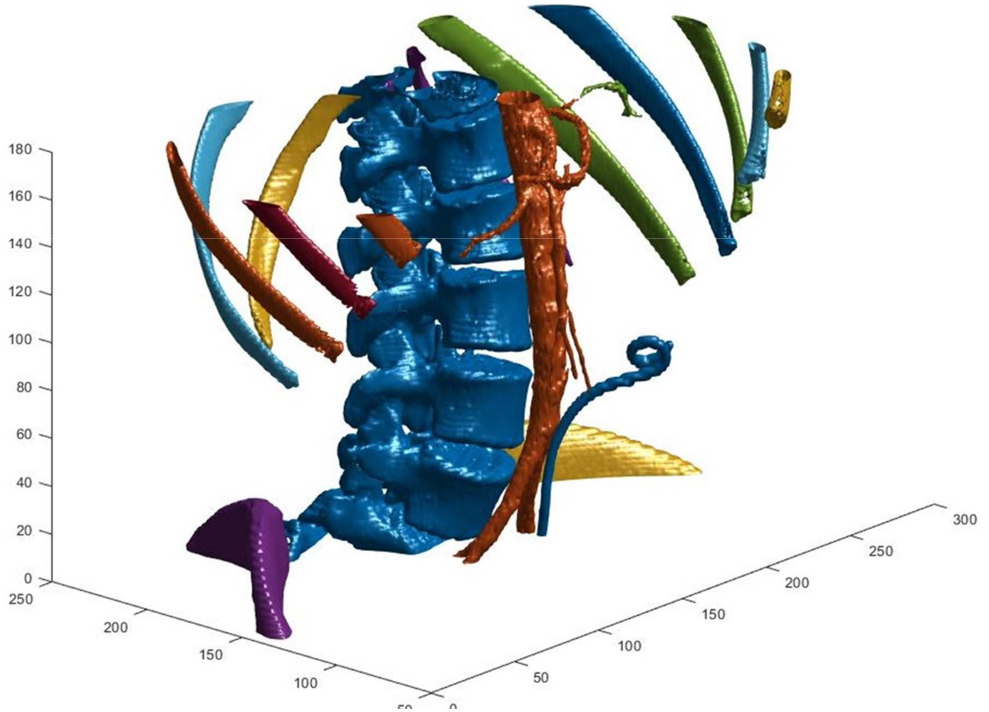
Segmentation example 1, abdominal CT. Segmentation of more dense objects splits ribs, backbone, aorta (very dense walls), beginning of hip and even a metallic prosthesis.

**Fig. 4.**
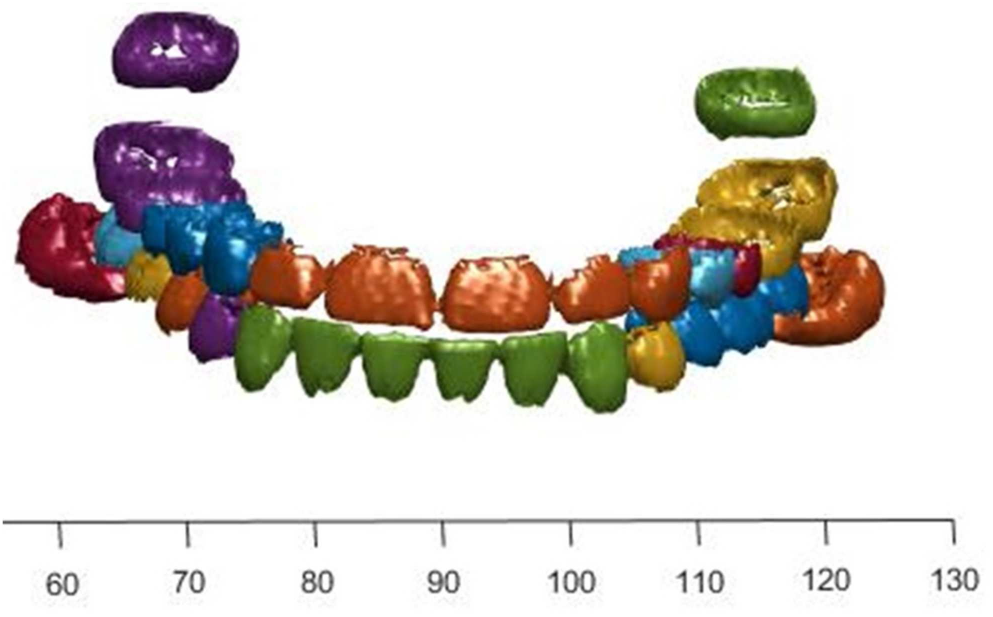
Segmentation example2, dental CT. Segmentation of more dense objects extracts teeth. Touching dental pieces are recognized as a single object.

For the case of MRI images, we did not obtain so good results. MRI’s are less resolution and less contrast images. Applying our methods directly does not get good segmentations as different tissues get connected. For the especial case of segmenting brain tissue in head MRI’s, we did a preprocessing applying a morphological filter: opening [12] to the individual slices before the isosurface stage (which permits to get rid of touching objects and select the biggest area object that, in this case, corresponds to brain). With this method, we can obtain a 3D object for the brain that has some defects due to the limited resolution (see figure 5).

**Fig. 5.**
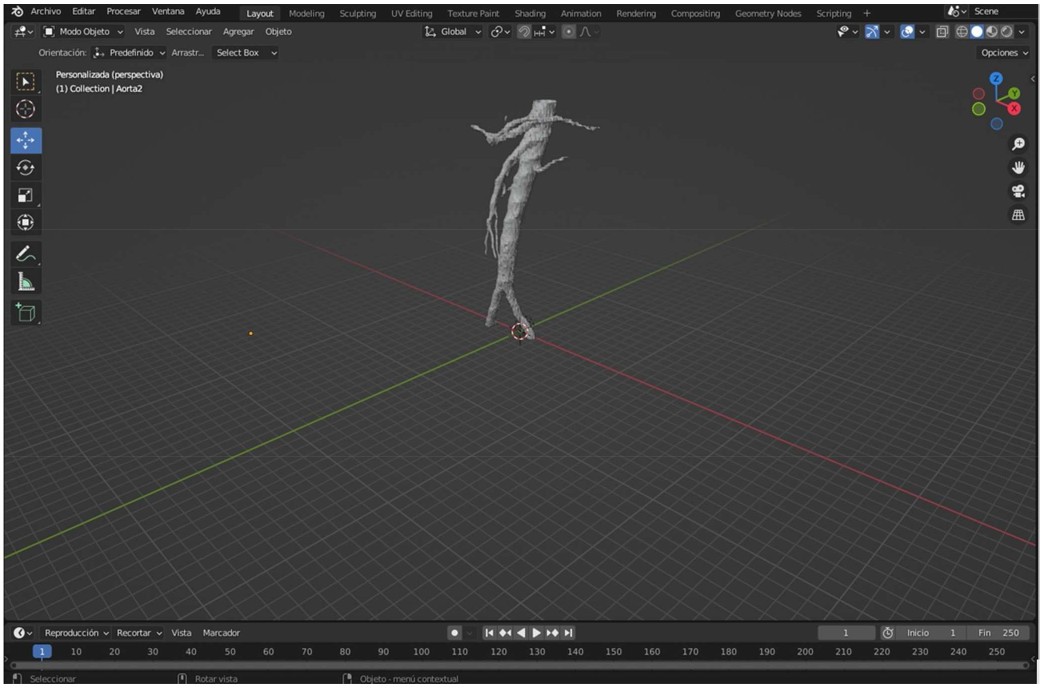
Aorta from figure 3 in Blender.

**Fig. 6.**
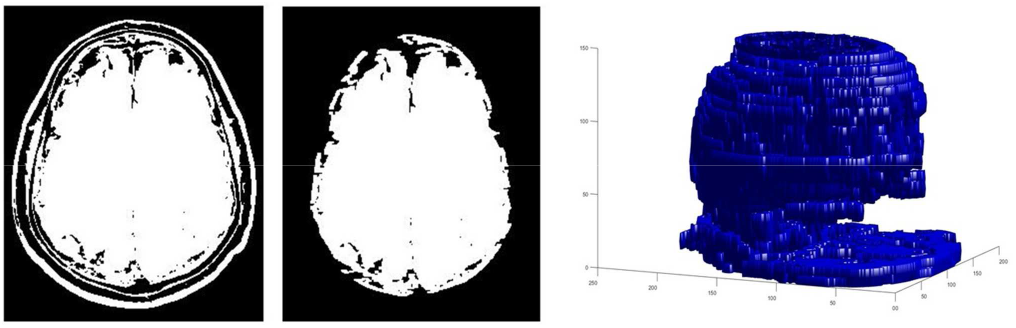
Brain 3D model from a MRI study.

## 4. Discussion

As main conclusion Gray Level threshold segmentation is good for CT images but is not appropriate for MRI.

As we think we have arrived to the limits of threshold techniques, future lines must be about exploring other algorithms and trying to integrate them with the present development.

Revising other methods described in [4], we now will describe them briefly trying to identify possible strategies for integration with ours:

- <u>Region Growing</u>: this method needs a pre-calculated set of initial points for each object (seeds). From the seeds, regions are built growing according to certain similarity criteria (than can be, but not limited to, similar gray level). Region growing is more versatile than thresholding but it depends much on previous knowledge: number of objects, seeds… Integration could come from using thresholding to get some of that previous knowledge.
- <u>Watershed</u>: watershed is a kind of region growing where image is interpreted as a “topographic” map where gray level is altitude. Seeds are “sources” of water, and computed contours are the points were water from different sources encounter. At this “frontiers”, dividing lines (watersheds) are built. Again this method is dependent from “a priori” knowledge. Sources must be local minima but using all of them would produce over-segmentation. Again a method based on histogram can help to get previous insight.
- <u>Level Set</u>: this method is more complex and it is based on a differential equation that allows to compute a surface that grows adaptively until being the segmented solution. The threshold method can be used to create an initial surface so that Level Set is faster and more precise.

Note that we have only found 2D implementations of these alternative methods (that operate slice by slice) where as we have developed a 3D one. What’s more watershed is intrinsically a 2D method. Creating a 3D version of region growing seems straightforward but it is not the same case for Level Set.

## Data Availability

All data produced in the present study are available upon reasonable request to the authors

## Acknowledgments

Authors have received no funding for this research. Authors acknowledge their institutions: University of Vigo and AtlantTIC for their support and for the necessary resources supplied.

